# Machine Learning-Driven Decision-Support System for Nursing Risk Assessment in Post-Discharge Care: A Design Science Approach

**DOI:** 10.1101/2025.10.20.25338376

**Authors:** Matilde M. Santos, Mariana Peyroteo, Luís V. Lapão

## Abstract

Hospital readmissions represent a persistent challenge for healthcare systems, often stemming from inadequate post-discharge monitoring. This study presents a Machine Learning (ML)-driven Clinical Decision Support System (CDSS) designed to enhance nursing risk assessment in post-discharge care.

Developed using a Design Science Research Methodology, the artefact integrates a digital questionnaire, an ML-based risk stratification model, and a real-time dashboard to optimise follow-up processes. The system was developed at a medical-surgical inpatient unit of a private hospital in Portugal. A retrospective dataset of 10,134 structured telephone follow-up records, classified by nurses into three risk levels—stable (A), requiring reassessment (B), and clinically concerning (C)—was used to train and evaluate the ML model.

Among the evaluated classifiers, Logistic Regression was selected for deployment based on its high specificity (0.9993), precision (0.9879), and absence of critical false negatives, despite slightly lower recall compared to XGBoost. The CDSS enables real-time risk classification based on patient-reported outcomes, supporting timely identification and prioritisation of patients requiring clinical attention. Simulation results indicate a potential reduction of up to 79% in nurse follow-up workload, while preserving care quality by focusing resources on moderate- and high-risk patients. Although direct evidence of reduced readmissions is not yet available, the system aligns with established best practices in transitional care. This study demonstrates the feasibility and utility of ML-based dynamic risk stratification for post-discharge monitoring, offering a scalable and interpretable solution that enhances clinical decision-making and resource allocation in nursing practice.

## 1. Introduction

### 1.1. Hospital Readmissions: Context and Impact

Hospital readmissions continue to represent a significant clinical and economic challenge for healthcare systems worldwide [2, 3]. Defined as patient re-hospitalisations occurring within a specific timeframe — typically 30 days post-discharge [4, 1] — they are associated with increased morbidity, lower patient satisfaction, and substantial financial burden to healthcare institutions [2, 3, 27]. In Europe, readmission rates range from 5% to 29% [2, 3, 1, 7, 5], with similar figures observed in Portugal, where general medicine readmissions within 30 days have been reported at 8.8% [1].

The principal causes of hospital readmissions include the progression of chronic conditions, inadequate discharge planning, and insufficient post-discharge follow-up [5]. Notably, deficiencies in discharge procedures—such as limited patient education, poor inter-professional communication, and absence of structured post-discharge support—are directly linked to increased readmission rates [5]. Evidence supports that identifying high-risk patients before discharge and ensuring coordinated follow-up care can reduce the likelihood of readmissions [6].

### 1.2. Post-Discharge Monitoring

Post-discharge monitoring aims to detect early signs of patient deterioration, promote treatment adherence, and provide reassurance during the recovery period [7]. Standard approaches include scheduled follow-up appointments and telephonic follow-up (TFU), typically conducted by nurses [6, 9]. TFU has demonstrated positive outcomes in reinforcing discharge instructions, enhancing medication compliance, and improving patient satisfaction [7, 9, 10, 26].

Despite these benefits, TFU implementation faces multiple operational challenges. These include invalid contact information, low response rates, delayed discharge notifications, and high workload among nursing staff, making the process time-consuming and resource-intensive [5, 9, 8, 11]. Such limitations highlight the need for innovative, scalable, and automated solutions to support effective post-discharge care.

### 1.3. Risk Assessment

Risk assessment is a fundamental process in healthcare, encompassing both risk prediction estimating the likelihood of future adverse events [16, 18] — and risk stratification categorising patients based on urgency or case severity to prioritise care [16, 14]. These approaches are complementary and context-specific [14, 16, 15].

In post-discharge settings, risk prediction is commonly operationalised through tools such as the LACE Index and the HOSPITAL Score, which evaluate demographic and clinical parameters to estimate the probability of 30-day readmission [18, 12, 13]. However, these tools have shown limited performance across diverse populations, with AUROC values ranging from 0.58 to 0.75 [13, 12]. Risk stratification, in contrast, is frequently applied in pre-admission scenarios such as emergency departments and telephone triage [21, 20], where tools like the Manchester Triage System support timely decision-making based on presenting symptoms. These methods allow for targeted care delivery but are rarely extended to post-discharge follow-up [19, 21].

### 1.4. Machine Learning-Driven Decision Support Systems

Recent advancements in Machine Learning (ML) have facilitated the development of data-driven models capable of improving both risk prediction and stratification [16, 17, 28]. ML algorithms such as Random Forest (RF), Logistic Regression (LR), and eXtreme Gardient Boosting (XGBoost) have demonstrated superior performance over traditional models in predicting adverse outcomes including readmissions, particularly when incorporating Electronic Health Record (EHR) data [28].

However, ML adoption in post-discharge settings remains limited. A recent scoping review revealed that most studies rely on static in-hospital data, with minimal integration of post-discharge patient-reported outcomes [28]. Moreover, while ML-based risk stratification has been increasingly explored in pre-admission care [21], its application to dynamically adjust post-discharge follow-up intensity remains underdeveloped.

### 1.5. Goal of the Study

This study aims to design, develop and evaluate an ML-driven Clinical Decision Support System to enhance nursing risk assessment in post-discharge care. Addressing limitations of static, discharge-based models, the proposed system incorporates patient-reported outcomes collected after discharge to dynamically stratify risk and guide follow-up necessities. The risk stratification is performed by a supervised ML model trained to replicate nurse-assigned classifications following telephone follow-up.

## 2. Materials

### 2.1. Healthcare Setting

The study was conducted at the Medical-Surgical Inpatient Unit of a private hospital in the Lisbon region, Portugal. This unit admits adult patients undergoing surgical procedures requiring overnight hospitalisation, as well as those admitted under internal medicine for acute conditions or diagnostic evaluation. It comprises 65 beds with an average occupancy rate of 70%, and an average patient age of 62 years old.

The multidisciplinary team includes internal medicine physicians, surgeons, and 20 nurses working in shifts, five of whom are responsible for conducting post-discharge TFU calls. The unit follows a structured nurse-led TFU programme, as part of its post-discharge care strategy. This setting provided a relevant context for assessing existing limitations and developing a data-driven solution to support clinical decision-making.

### 2.2. Database

A retrospective, anonymised clinical dataset was used, derived from the nurse-led TFU programme. Due to institutional restrictions, the dataset is not publicly available. Data were collected by nurses during structured follow-up calls and initially comprised 53 variables: eight related to administrative and management processes, 42 reflecting patient-reported health status during the call, and three concerning patient satisfaction.

The study cohort included all patients admitted between 1 January 2021 and 31 December 2023 who received at least one documented follow-up call. Of the 12,879 records retrieved, those lacking a nurse-assigned classification were excluded, as they represented call attempts rather than completed assessments. This yielded a final dataset of 10,134 valid records, with each entry corresponding to a single follow-up call.

For ML model development, only the 42 variables concerning health status were retained. Satisfaction-related variables were excluded due to their collection after the nurse’s classification, to avoid introducing post-outcome bias. Administrative and management variables were also removed, resulting in a final modelling dataset with 42 variables per record.

The model was developed as a supervised multi-class classifier trained to replicate the nurse-assigned classification. The target variable was the nurse-assigned classification, categorised as A (clinically stable), B (requiring reassessment), and C (clinically concerning), with a highly imbalanced distribution: 9987 (A), 59 (B), and 88 (C).

### 2.3. Research Team

The study was developed by a multidisciplinary team of five members: three researchers from NOVA University of Lisbon with expertise in biomedical engineering and digital health, and two clinicians from the hospital, including the unit coordinator and a nurse. This collaboration ensured alignment between technical development and clinical practice throughout the study.

## 3. Methods

### 3.1. Study Design

The research followed the Design Science Research Methodology (DSRM), a structured approach suited for developing artefacts that address real-world problems, particularly in complex healthcare settings [22, 23]. DSRM comprises six iterative stages: (1) Problem Identification, (2) Definition of Objectives, (3) Design and Development, (4) Demonstration, (5) Evaluation, and (6) Communication [22]. Each stage was executed as a research activity and validated in collaboration with the clinical team.

### 3.2. Problem Identification

The objective of this phase was to identify the limitations of the current post-discharge follow-up process. The process mapping was developed using three complementary methods: process observations, multidisciplinary meetings, and Exploratory Data Analysis (EDA) of the clinical dataset. A shadowing approach was adopted to observe 21 follow-up calls conducted by three different nurses, enabling real-time monitoring of care delivery and workload management with minimal disruption to clinical routines. Additionally, a structured set of meetings with the clinical team were conducted to map the steps involved in the TFU process and to identify operational challenges. EDA of the TFU dataset complemented these findings, allowing identification of operational bottlenecks and inconsistencies in classification and symptom reporting patterns.

### 3.3. Objectives for a Solution

Based on the process mapping and insights gained during the previous phase, this activity aimed to define the objectives of the proposed artefact and establish a revised follow-up process. To achieve this, insights from the previous phase were analysed collaboratively with the clinical team.

### 3.4. Design and Development

This phase involved the creation of the three main components of the CDSS artefact: the ML model, the data acquisition mechanism, and the graphical visualisation interface. This stage was developed in close collaboration with the clinical team, with often validations of the CDSS artefact.

#### 3.4.1. ML Model

A supervised ML model was developed to replicate nurse-assigned classifications during follow-up calls. The complete pipeline for data preparation, modelling, and evaluation is illustrated in Figure 1.

**Figure 1:**
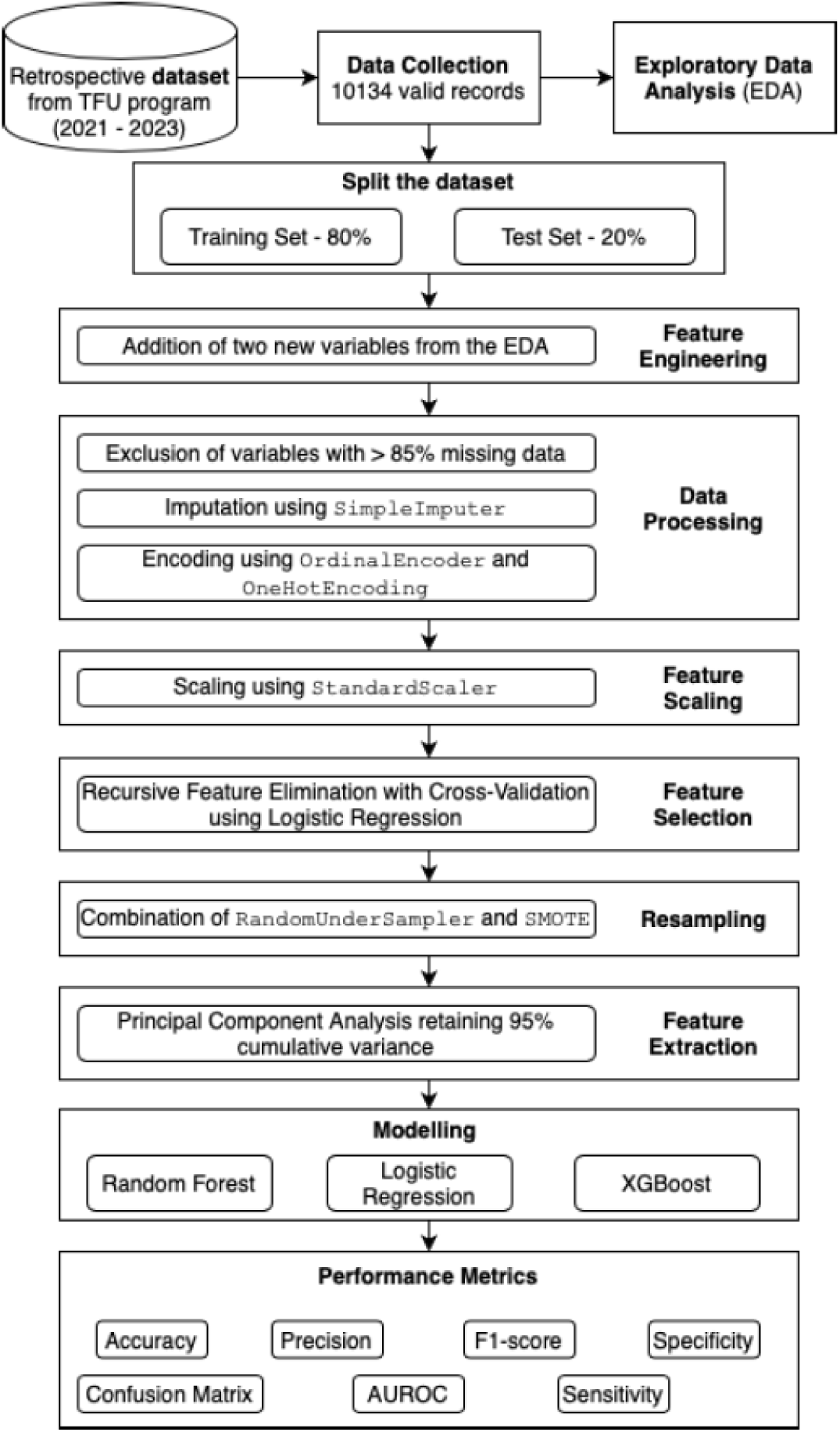
Pipeline of the ML risk stratification model

Two new variables (“Had a recent medical appointment or one coming up?” and “Weighted Total of Symptoms”) were derived based on insights from the EDA. Before any processing, the dataset was split using a stratified sampling strategy into a training set (80%) and a test set (20%) to preserve class proportions.

During the data processing stage, variables with more than 85% missing data were excluded. For those with clinically meaningful missingness, values were imputed using the SimpleImputer class from sklearn with a constant strategy [25]; the remaining missing values were imputed using the most frequent category, as all variables were categorical. Encoding of categorical variables was performed using the class OrdinalEncoder for ordinal features and the class OneHotEncoder for nominal ones. All features were standardised using the class StandardScaler.

Feature selection was performed using Recursive Feature Elimination with Cross-validation with LR as the base estimator and the class StratifiedKFold for cross-validation. Data imbalance was addressed by combining the class RandomUnderSampler with Synthetic Minority Over-Sampling Technique.

Dimensionality reduction was performed using Principal Components Analysis, retaining components accounting for 95% of cumulative variance.

Three models were trained — RF,LR, and XGBoost — and tuned using the class GridSearchCV with 5-fold stratified cross validation. Evaluation metrics included accuracy, precision, sensitivity, specificity, F1 score, AUROC, and normalised confusion matrices.

#### 3.4.2. Data Acquisition

A digital questionnaire was developed using Google Forms, to replace the initial follow-up call and enable automated risk assessment. The questionnaire was co-designed with the clinical team to ensure consistency with the existing TFU protocol. Responses were stored in a Google Sheets database, allowing real-time access by the ML model.

#### 3.4.3. Graphical Visualisation

A web-based dashboard was developed using Streamlit to support clinical decision-making. The interface enables nursing staff to view risk classifications, prioritise follow-up actions, and access patient-reported data in a structured and efficient format.

### 3.5. Demonstration

The three system components—ML model, digital questionnaire, and dashboard—were integrated into a functional artefact. Google Sheets was used as a central data repository, enabling real-time synchronization between the questionnaire, model, and dashboard. The system leverages the job lib library for model deployment and the Google Sheets API for real-time data retrieval and update.

### 3.6. Evaluation

The evaluation assessed whether the developed artefact met the previous defined objectives. The system’s potential impact was estimated based on projected reductions in follow-up workload and improved prioritisation of high-risk patients. Feedback from the clinical team during a structured workshop was used to validate usability, clinical relevance, and alignment with workflow needs.

### 3.7. Communication

Communication activities were conducted throughout the research to ensure alignment with clinical needs and promote the dissemination of the findings. Regular meetings with the clinical team supported iterative development and validation. Results were also shared through a multidisciplinary workshop and the submission of peer-reviewed publications.

### 3.8. Ethical Consideration

Ethical approval was granted by the hospital’s Administration and Clinical Board and its Ethics Committee for Health. All data were anonymised before analysis and managed in accordance with the General Data Protection Regulation (GDPR).

## 4. Results

### 4.1. Problem Identification

The initial phase of the study focused on identifying inefficiencies within the post-discharge follow-up process currently implemented in the Medical-Surgical Inpatient Unit. A combination of direct observation, multidisciplinary meetings, and EDA was used to map the workflow and assess its effectiveness.

The mapped process revealed a uniform follow-up approach, where all discharged patients received a telephone call irrespective of their clinical status or perceived risk. Observational data highlighted variability in practice and workload across the nursing team, while meetings with clinical stakeholders confirmed recurrent operational constraints. These included excessive time demands, duplicated tasks, and lack of prioritisation criteria.

The EDA of the clinical dataset supported these findings, showing that most patients (98.6%) were classified as low risk, suggesting that a large portion of follow-up calls may be of limited clinical value (i.e., contributing to inefficiency). Additionally, inconsistencies were identified in the documentation of symptoms and risk classifications, further emphasising the need for standardisation and decision support.

Overall, the process was deemed inefficient due to its disproportionate resource consumption and ineffective as it failed to identify and prioritise higher-risk patients. Furthermore, the absence of continuous monitoring mechanisms limited the system’s responsiveness to evolving patient needs following discharge.

### 4.2. Objectives of the Solution

In response to the identified limitations, two primary objectives were defined by the research and clinical team for the proposed solution:

1. To improve the efficiency of the follow-up process by reducing the time nurses dedicate to follow-up activities.
2. To enhance effectiveness by enabling the timely identification of patients at risk of readmission through targeted, risk-based interventions.

A revised workflow was proposed, integrating a CDSS that uses an MLdriven risk stratification model (see **Appendix A**). In the new process, only patients classified as moderate or high risk (yellow or red) would receive telephone follow-up, allowing nursing resources to be focused where most clinically needed. Low-risk patients (green) would be monitored digitally, thus reducing manual workload while maintaining continuity of care.

### 4.3. Design and Development

#### 4.3.1. ML Risk Stratification Model

Table 1 presents the performance metrics for three models: RF, LR, and XGBoost, with the highest values in each category highlighted. XGBoost achieved the best results in accuracy (0.9206), sensitivity (0.9206), and F1 score (0.9490), indicating strong overall performance and effective identification of high-risk patients. In contrast, LR outperformed the other models in precision (0.9879), specificity (0.9994), and AUROC (0.9879), suggesting superior capability in minimizing false positives and enhancing overall discriminative power. While RF showed competitive results, it did not lead in any metric.

**Table 1:** Performance metrics for each model, with the best values highlighted in blue.

| Model | Accuracy | Precision | Sensitivity | F1 score | Specificity | AUROC |
| --- | --- | --- | --- | --- | --- | --- |
| RF | 0.9038 | 0.9873 | 0.9038 | 0.9399 | 0.9665 | 0.9842 |
| LR | 0.9068 | 0.9879 | 0.9068 | 0.9416 | 0.9994 | 0.9879 |
| XGBoost | 0.9206 | 0.9863 | 0.9206 | 0.9490 | 0.9010 | 0.9841 |

Further insights are provided by the normalized confusion matrices for LR and XGBoost (see Figure 2). LR showed higher classification consistency for class B (0.92) and lower misclassification rates overall, particularly for class C, where 83% of cases were correctly classified and 17% misclassified as B. XGBoost demonstrated slightly higher performance for class A (0.92) and class C (0.83), but a lower classification rate for class B (0.75), with 17% of B instances misclassified as class C.

**Figure 2:**
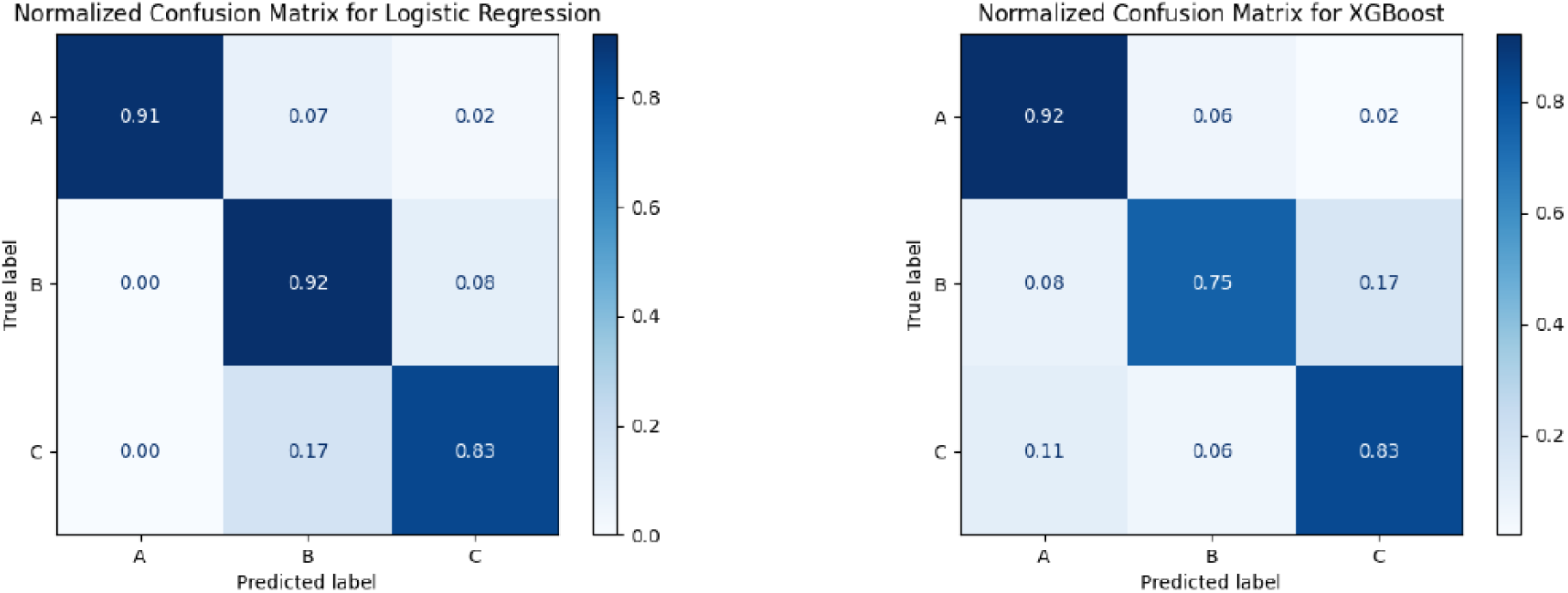
Normalized Confusion Matrices of LR and XGBoost.

These findings highlight the complementary strengths of XGBoost and LR, each excelling in different performance dimensions. XGBoost is particularly suitable for scenarios prioritizing sensitivity, whereas LR may be preferred when specificity and interpretability are critical. As no single model consistently outperforms across all metrics, model selection should be guided by clinical priorities and the specific trade-off required in the healthcare setting.

#### 4.3.2. Clinical Decision Support System

The questionnaire was structured into five sections and designed to be user-friendly, avoiding technical medical terminology. It includes mandatory questions identified through feature selection, complemented by free-text fields to provide additional context to the nursing team. The form features brief, plain-language introductions to each section, guiding patients through the process.

Developed in Portuguese, it ensures linguistic and cultural appropriateness for the hospital’s patient population. The questionnaire content aligns directly with the model’s selected predictive variables and supports real-time risk classification.

The dashboard, also developed in Portuguese and co-designed with the clinical team, presents real-time risk predictions and facilitates structured follow-up. Upon login, users access three main interface views. The Home Screen displays each nurse’s agenda, including scheduled and overdue calls, with summaries of high- and moderate-risk patients and a progress bar tracking completed calls. The Call Management Screen lists patients requiring follow-up, prioritised by urgency. Nurses can view responses, register calls, and assign classifications. Green patients are removed; yellow and red remain until follow-up is complete. After three unanswered attempts, patients can be excluded. The Complete Patient List provides a searchable, filterable history of all follow-up records.

### 4.4. Demonstration

The fully functional CDSS was demonstrated through the successful integration of its key components: the post-discharge questionnaire, the risk stratification model, the shared database, and the clinical dashboard. These elements were connected to enable real-time classification and follow-up decision support, forming a complete digital artefact for post-discharge care.

Figure 3 illustrates the information flow within the system. Patients initiate the process by completing the post-discharge questionnaire via Google Forms. Their responses are automatically recorded in a centralised database implemented using Google Sheets (step a). The ML-based risk stratification model then retrieves this input (step b), processes the data, and updates the database with a risk classification (step c).

**Figure 3:**
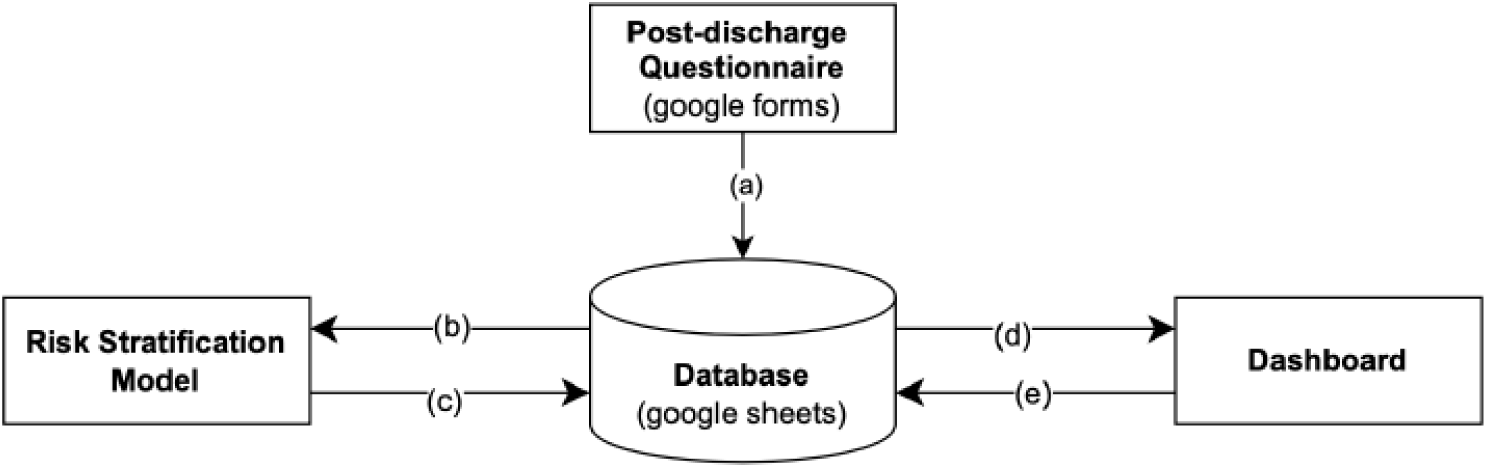
Information Flow in the iFollow@Care artefact.

The dashboard, accessed by the nursing team, reads the updated classifications from the same database (step d), enabling immediate visualisation of patient status and follow-up priorities. Any actions performed by nurses, such as logging call outcomes, assigning classifications, or updating patient information, are written back to the database (step e), ensuring synchronisation across all components in real-time.

### 4.5. Evaluation

The redesigned follow-up process introduces a risk-based approach that prioritises only moderate- and high-risk patients (classified as yellow and red) for telephone contact. Patients identified as low-risk (green) are automatically excluded from manual follow-up, enabling a substantial reduction in nursing workload. Data collected indicated that 98.6% of patients were classified as green. However, considering evidence from the literature suggesting that 12–19% of patients may over-report symptoms [29], a conservative estimate was applied, projecting a maximum achievable reduction of approximately 79% in follow-up volume.

In operational terms, nurses currently perform an average of 65 follow-up calls per week, each lasting around seven minutes. This corresponds to a total weekly follow-up time of approximately 455 minutes (7 hours and 35 minutes). Under the revised process, the expected reduction in workload would lower this to approximately 96 minutes per week (1 hour and 36 minutes).

The second objective relates to the potential impact of the artefact on reducing 30-day hospital readmissions. While direct evidence of reduced readmissions was not yet measurable within the scope of this study, the literature consistently highlights the importance of structured post-discharge monitoring in preventing avoidable readmissions. The implementation of a dynamic, risk-based follow-up process, combined with the possibility of extending monitoring to 30 days, aligns with best practices in transitional care and offers a strong foundation for improving long-term outcomes.

## 5. Discussion

### 5.1. Selection of the best model according to the clinical healthcare setting

The selection of the optimal machine learning model in this study was guided by clinical priorities rather than by overall performance metrics alone. In this setting, the primary objective is to avoid misclassifying patients who require follow-up—those assessed as moderate- or high-risk (B or C)—as clinically stable (A). These false negatives are the most serious type of classification error, as they result in patients being excluded from the follow-up process, potentially leaving them without the necessary medical attention.

As shown in the confusion matrices (see Figure 2), LR demonstrated the strongest performance in this regard, avoiding all false negatives involving moderate- and high-risk patients misclassified as low-risk. This guarantees that all patients requiring follow-up would be flagged for clinical review. In contrast, XGBoost misclassified 8% of moderate-risk and 11% of high-risk patients as stable, introducing a significant risk of missed interventions.

A secondary concern involves the misclassification of high-risk patients as moderate-risk. Although these patients would still receive follow-up, the delay in intervention could compromise clinical outcomes. In this error category, XGBoost showed stronger performance, misclassifying only 6% of high-risk patients as moderate risk, compared to 17% under LR.

False positives, where stable patients are mistakenly classified as moderate or high-risk, pose less clinical risk but increase nursing workload due to unnecessary follow-ups. LR generated fewer of these cases, making it more efficient in terms of resource allocation.

These findings indicate that while both models have strengths, LR aligns more closely with the clinical requirements of this healthcare setting. Its ability to avoid the most critical classification errors, combined with its efficiency in minimising unnecessary interventions, makes it the most appropriate model for deployment within the developed CDSS.

### 5.2. Comparative Analysis with Existing Models

The selected LR model was evaluated against machine learning approaches reported in recent literature, with particular reference to a recent scoping review on post-discharge solutions with AI [28]. This review summarised a broad range of model performances in post-discharge care, reporting AUROC values between 0.6950 and 0.9940 across diverse settings.

The LR model developed in this study achieved an AUROC of 0.9879, placing it near the upper bound of reported values. Its recall of 0.9068 also compares favourably, falling within the range of 0.6500 to 0.9470 observed in the literature. In addition to these core performance metrics, the model demonstrated exceptional specificity (0.9993) and precision (0.9879), both exceeding the highest values reported in the review (0.9130 and 0.9510, respectively). These results suggest a low rate of false positives and a high degree of reliability in identifying patients requiring follow-up.

Other studies have predominantly focused on models trained with retrospective clinical data extracted from EHR, often limited to information available at the time of discharge [28, 17]. These models, while effective in certain contexts, do not account for changes in a patient’ s condition during the post-discharge period and rarely incorporate patient-reported outcomes. As a result, their applicability to dynamic follow-up processes is constrained.

In contrast, the LR model in this study was designed to operate with structured, patient-reported data and developed in collaboration with clinical staff. Its combination of high predictive performance, low complexity, and strong clinical interpretability makes it particularly suitable for integration into nurse-led follow-up processes. Compared to models reported in the literature, it offers a competitive and practical solution for post-discharge risk stratification.

### 5.3. Comparative Analysis with traditional risk score models

Traditional risk prediction tools such as the LACE Index and HOSPITAL Score have shown limited discriminative performance, with AUROC values typically ranging between 0.58 and 0.75 [13, 12]. These models rely on static discharge data and do not account for the patient’s evolving condition after hospitalisation [18, 13, 12].

In contrast, the LR model developed in this study achieved an AUROC of 0.9879, demonstrating significantly stronger predictive accuracy. Unlike traditional tools, the model incorporates patient-reported outcomes collected post-discharge, enabling dynamic risk assessment.

This approach not only improves performance but also aligns more closely with clinical needs by supporting timely, risk-based follow-up.

### 5.4. Assessment of the artefact using the Österle’s Design Principles

The developed artefact was evaluated through the lens of the four Österle principles — abstraction, originality, justification, and benefits [24]—which serve as a theoretical foundation for assessing contributions in design-oriented information systems research. Two hospital physicians, two nurses, an engineer specialised in health information systems, and a digital health expert participated in the assessment. This framework supports a critical discussion of the artefact’s broader value and relevance.

#### Abstraction

While designed for post-discharge follow-up in a medical-surgical inpatient context, the artefact addresses a more general problem class related to risk-based patient monitoring beyond hospital discharge. Its underlying logic, using self-reported data to dynamically stratify patients by risk, canbe adapted to other use cases such as post-procedure recovery or medication monitoring. This abstraction potential strengthens the artefact’s relevance across diverse clinical scenarios.

#### Originality

The solution introduces a novel approach by combining patient-reported outcomes with machine learning for real-time clinical risk stratification. Existing models in the literature typically rely on retrospective hospital data or static discharge metrics, whereas this artefact integrates timely, subjective patient data into a dynamic decision-making process. No comparable systems were identified that implement this logic for follow-up care, indicating a meaningful contribution to the field of digital health innovation.

#### Justification

The artefact was developed in response to clearly defined clinical challenges, including inefficient resource allocation and the lack of prioritisation in standardised follow-up protocols. The system directly addresses the problem it was designed to solve through the early identification of higher-risk patients and the automated exclusion of low-risk cases. Its alignment with clinical workflows further supports its practical justification.

#### Benefits

The artefact supports measurable improvements in both efficiency and effectiveness. The reduction in unnecessary follow-up calls enables more efficient allocation of nursing resources. At the same time, it fosters a more proactive and personalised approach to patient monitoring. Additionally, the system promotes digital literacy among healthcare professionals and supports greater patient engagement in post-discharge care, contributing to broader institutional and behavioural benefits.

The artefact meets all four Österle principles, offering a transferable, innovative, problem-driven, and benefit-oriented contribution. These findings reinforce its value as both a practical solution and a research outcome in the domain of digital health.

### 5.5. Limitations of the study

This study presents some limitations that should be acknowledged when interpreting the results. First, the dataset used did not include demographic variables such as age, sex, or socioeconomic status. Although these factors are known to influence post-discharge outcomes, they were not available from the healthcare provider and could not be incorporated into the risk model.

Second, the study lacked access to hospital readmission data, which limited the ability to directly assess the system’s impact on reducing 30-day readmissions. While the model’s performance suggests strong potential for effective follow-up risk stratification, its influence on actual readmission rates remains to be validated.

Third, the model evaluation was restricted to three machine learning algorithms: LR, RF, and XGBoost. Although these models performed well, future advances in artificial intelligence may offer alternative techniques better suited to this application.

Lastly, the system was validated using retrospective real-world data but not yet implemented in an on-site clinical environment. As such, its practical impact on workflow integration, user experience, and clinical outcomes will require further investigation through prospective on-site deployment.

### 5.6. Future Work

Future work will focus on addressing the limitations identified in this study and advancing the system’s functionality to support broader clinical integration. A key area for development is the integration of the platform with EHR. Access to additional patient data, particularly demographic variables such as age, sex and socioeconomic status, would enable a more comprehensive risk assessment and improve the model’s predictive accuracy. Incorporating these factors into the ML model is essential to enhance its generalisability and clinical relevance.

Another area of focus is ensuring transparency and trust in algorithm supported decision-making. Although the system currently flags low-risk patients for exclusion from follow-up, a validation step could be introduced whereby nurses review and confirm all low-risk classifications before they are finalised. This would reinforce the clinician’s central role in the process and promote confidence in the system’s outputs.

Patient communication also presents an opportunity for enhancement. Currently, low-risk patients do not receive direct feedback after completing the questionnaire. Implementing an automated message to reassure patients that their condition has been reviewed and appears stable, along with providing a contact point for follow-up concerns, may improve engagement and perceived continuity of care.

From a technical perspective, this study evaluated three machine learning models. Future research should explore the potential of deep learning and other advanced ML techniques to further improve risk stratification performance, particularly in cases with more complex data patterns.

Finally, an on-site clinical pilot is already planned to assess the system’s practical impact on workflow efficiency, user experience, and patient outcomes, including its potential effect on 30-day readmission rates.

In the longer term, the platform could also evolve into a conversational agent-based system, leveraging chatbot interfaces to streamline patient interaction and reduce clinician workload, while maintaining the structured follow-up approach established in this study. Also, adding robotic process automation could make the system even more efficient, taking care of repetitive tasks like automatically sending follow-up forms, issuing reminders, and updating the patients as soon as responses are reviewed.

## 6. Conclusions

This study presented the design, development, and evaluation of a machine learning-driven CDSS to support nursing risk assessment in post-discharge care. Developed using a Design Science Research approach, the artefact integrates patient-reported outcomes, an ML-based risk stratification model, and a real-time dashboard to support follow-up risk stratification and resource optimisation.

The selected LR model demonstrated strong predictive performance, outperforming both traditional scores (LACE and HOSPITAL) and several MLbased approaches reported in the literature. Its use of post-discharge data and high specificity and precision render it particularly suitable for real-world deployment in nurse-led follow-up programmes.

The system achieved its intended objectives by significantly reducing unnecessary follow-up workload— with an expected optimisation of up to 79%— while supporting the timely identification of high-risk patients. It also promotes digital engagement among healthcare professionals and patients, aligning with broader goals in digital health innovation.

While limitations exist, particularly regarding the absence of demographic and readmission data, and the lack of on-site implementation, this work establishes a robust foundation for future use of artificial intelligence in clinical integration and improving workforce efficiency. The proposed system offers a scalable, interpretable, and clinically aligned solution for enhancing post-discharge monitoring through data-driven decision support.

## Data Availability

All data produced in the present study are available upon reasonable request to the authors

## 7. Acknowledgements

The authors would like to express their sincere gratitude to Dr Marta Jonet, Nurse Rita Tinoco, and Nurse Carolina Barca for their invaluable contributions to the development of this project. Their clinical insight, commitment, and support were instrumental in ensuring the system’s relevance and integration into nursing practice. The authors also acknowledge the Lusíadas Amadora Hospital for providing the necessary institutional support and access to clinical data, which were essential to the successful execution of this study. The authors further thank the hospital’s Ethics Committee for approving the study and supporting its development. Authors acknowledge Fundação para a Ciência e a Tecnologia I.P. for its financial support via the project <u>UID/00667</u>: Unidade de Investigação e Desenvolvimento em Engenharia Mecânica e Industrial. MP was supported by FCT (<u>2023.00994.BD</u>). In both cases, the funding body provided financial support for the research, with no involvement in the analysis or writing of the article.

## Ethics Statement

Ethical approval for the study was granted by the Administration and Clinical Directorate of Hospital Lusíadas Amadora, as well as the Ethics Committee for Health of Hospital Lusíadas Amadora. All patient data were anonymised before analysis and handled in accordance with the General Data Protection Regulation (GDPR). No individual-level consent was required as the dataset was fully anonymised and collected retrospectively from routine care.

## Appendix A. Proposal for Improved Follow-Up Process Framework

**Figure A.4:**
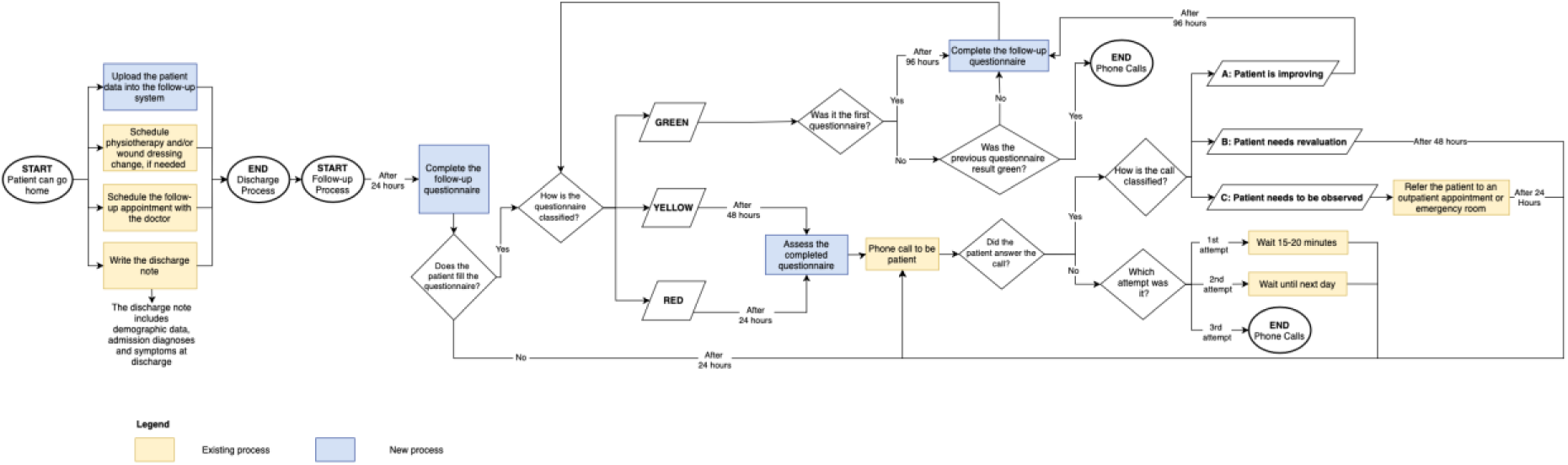
Proposal for Improved Follow-Up Process.

